# Alzheimer’s disease susceptibility gene apolipoprotein e (*APOE*) and blood biomarkers in UK Biobank (N=395,769)

**DOI:** 10.1101/2020.02.12.20021998

**Authors:** Amy C. Ferguson, Rachana Tank, Laura M. Lyall, Joey Ward, Carlos Celis-Morales, Rona Strawbridge, Frederick Ho, Christopher D. Whelan, Jason Gill, Paul Welsh, Jana J. Anderson, Patrick B. Mark, Daniel F. Mackay, Daniel J. Smith, Jill P. Pell, Jonathan Cavanagh, Naveed Sattar, Donald M. Lyall

## Abstract

**Background and objective:** Alzheimer’s disease (AD) is a neurodegenerative condition where the underlying aetiology is still unclear. Investigating the potential influence of apolipoprotein e (*APOE*), a major genetic risk factor, on common blood biomarkers could provide a greater understanding of the mechanisms of AD and dementia risk. Our objective was to conduct the largest (to date) single-protocol investigation of blood biomarkers in the context of *APOE* genotype, in UK Biobank.

**Methods:** After quality control and exclusions, data on 395,769 participants of White European ancestry were available for analysis. Linear regressions were used to test potential associations between *APOE* genotypes and biomarkers.

**Results:** Several biomarkers significantly associated with *APOE* e4 ‘risk’ and e2 ‘protective’ genotypes (vs. neutral e3/e3). Most associations supported previous data: for example, e4 genotype was associated with elevated low-density lipoprotein cholesterol (LDL) (standardized beta [b] = 0.150 standard deviations [SDs] per allele, p<0.001) and e2 with lower LDL (b = −0.456 SDs, p<0.001). There were however instances of associations found in unexpected directions: e.g. e4 and increased insulin-like growth factor (IGF-1) (standardized beta = 0.017, p<0.001) where lower levels have been previously suggested as an AD risk factor.

**Conclusions:** These findings highlight biomarker differences in non-demented people at genetic risk for dementia. The evidence here in supports previous hypotheses of involvement from cardiometabolic and neuroinflammatory pathways.

## Introduction

Alzheimer disease (AD) is the most common form of dementia and an important public health issue [1], hypothesized to be the result of interactions between genetic and environmental risk factors [2]. *APOE* e4 genotype is the largest common single genetic risk factor for AD and cognitive decline behind increasing age [3], with the e2 allele potentially protective [1]. The exact mechanisms by which *APOE* genotype influences brain ageing are unclear but probably due to pleiotropic pathways stemming from its core role in lipid metabolism [4].

Several studies have investigated serum biomarker differences between AD patients vs. healthy individuals in order to identify potential risk factors for AD, including low density lipoproteins (LDL) and insulin-like growth factor 1 (IGF-1) with sometimes conflicting results potentially due to methodological heterogeneity [5,6]. Many studies investigating serum levels in AD have focussed on specific biomarkers involved in β-amyloid precursor protein metabolism and phosphorylation [7]. There have been relatively few studies investigating a wide range of biomarkers in a “hypothesis-free” approach; and those studies which have done this appear to be limited by small sample size or have been cross-sectional in individuals with an extant diagnosis of dementia [8–10]. Relatively few biomarkers have been investigated in the context of AD-susceptibility gene *APOE*. Gaining a greater understanding of how *APOE* influences biomarker serum levels could be extremely beneficial: highlighting factors significantly associated with AD genetic risk could elucidate potential pathways involved in its development, pathophysiology and ultimately treatment [11].

In this study *APOE* genotype status was tested vs. a range of circulating serum blood biomarkers available for approximately 396,000 participants in UK Biobank. Two separate analyses were undertaken to investigate the influence of genotypic status on biomarker levels: differences per i) risk e4, or ii) protective e2 allele; each vs. neutral e3/e3 genotype. Further analyses were undertaken to investigate the associations in males and females separately due to *a priori* evidence for *APOE*-sex differences in AD pathophysiology [12]. To our knowledge, this is the first large-scale investigation of the relationship between *APOE* genotype status and a wide range of biomarkers in a population cohort.

## Methods

### Subjects

Over 502,000 UK residents aged 37-73 years were recruited to UK Biobank from 2006-2010. At one of 22 assessment centres across the UK, participants completed a range of phenotypic assessments and questionnaires, including genetic, urine and blood samples [13]. We focussed on participants with White British ancestry because there is evidence of different e4 frequencies across ethnicities[14].

### Ethical approval

This secondary-data analysis study was conducted under generic approval from the NHS National Research Ethics Service (approval letter dated 17th June 2011, ref 11/NW/0382). Written informed consent was obtained from all participants in the study (consent for research, by UK Biobank).

### Genotyping

UK Biobank participants were genotyped using Applied Biosystems UK BiLEVE Axiom array by Affymetrix and Applied Biosystems UK Biobank Axiom Array which share 95% marker content [13]. *APOE* e status was based on two single nucleotide polymorphisms (SNPs): rs7412 and rs429358. Stringent quality control and processing were applied to the data, detailed at http://www.ukbiobank.ac.uk/scientists-3/genetic-data and http://www.ukbiobank.ac.uk/wp-content/uploads/2014/04/UKBiobank_genotyping_QC_documentation-web.pdf.

### Biomarker collection and processing

Biomarker levels were analysed in UK Biobank from serum and packed red blood cell samples obtained from all UK Biobank participants at baseline [15]. Stringent quality controls were applied to the assays used measure biomarker levels, details of biomarker quality control, instrumentation and analysis methods are available at: https://biobank.ndph.ox.ac.uk/showcase/showcase/docs/biomarker_issues.pdf, https://biobank.ndph.ox.ac.uk/showcase/showcase/docs/serum_biochemistry.pdf, http://biobank.ndph.ox.ac.uk/showcase/showcase/docs/haematology.pdf, and http://www.ukbiobank.ac.uk/wp-content/uploads/2018/11/BCM023_ukb_biomarker_panel_website_v1.0-Aug-2015-edit-2018.pdf. Processing of very low levels of oestradiol and rheumatoid factor were recorded as “missing” (in the original data); these missing values were recoded conservatively as the square root of the minimum stated detectable value if individuals had data for a remaining biomarker [16] and were not coded as ‘no data returned’ or having unrecoverable aliquot problems.

### Dementia outcomes

We validated *APOE* genotype’s association with dementia/AD outcomes (in UK Biobank) as a check, having previously shown associations in expected directions with brain structure [3] and cognitive abilities [17]. Dementia and AD outcomes were generated using self-report, hospital admission and death record data, with hospital and death record data utilising International Classification of Diseases version 10 (ICD-10 codes). Individuals were designated as cases (“all-cause dementia” or “Alzheimer disease”) if they had indicated dementia or AD in either self-report, or through hospital or death records – derived by UK Biobank based on self-report, hospital admission and death reports [18]. Those coded as missing were designated as controls (i.e. did not self-report dementia/AD, and these diagnoses were not present in hospital/death records).

### Covariates

Participants self-reported their smoking history, and we collated past and current smokers into ‘ever’ (vs. never). Participants self-reported medication use for cholesterol, high blood pressure or insulin. We excluded those who did not know or preferred not to answer for these various items (<5%). Townsend deprivation indices were derived from postcode of residence [19]. This provides an area-based measure of socioeconomic deprivation derived from aggregated data on car ownership, household overcrowding, owner occupation and unemployment. Higher Townsend scores equate to higher levels of area-based socioeconomic deprivation. We additionally controlled for potential population stratification using UK Biobank-derived principal components (PCs) 1-5, and genotypic array[13]. Height was measured (Seca 202 stadiometer) and weight was measured to the nearest 0.1 kg (BC-418 MA body composition analyser; Tanita Corp). Body mass index (BMI) was derived from weight in kilograms divided by height in meters squared. All-cause cancer was derived based on self-report at baseline. Month of assessment was recorded by UK Biobank. We have previously described and derived an ‘any self-reported inflammatory condition’ variable[20].

### Statistical analyses

The linear regressions reported reflect average SD-changes per e4 or e2 allele vs. neutral e3/e3 genotype, i.e. e3/e3 vs. e3/e4 vs. e4/e4 (dose), and e2/e2 vs. e2/e3 vs. e3/e3 (dose). Associations with *APOE* genotype vs. dementia/AD were tested using binary logistic regressions reporting odds ratios (OR) and their 95% confidence intervals. We corrected for multiple testing using False Discovery Rate (FDR)[21], conservatively collating all tests. Biomarkers which were not normally distributed were log transformed prior to Z-score transformation and reanalysed: the resulting effect sizes were unchanged and therefore we report the original estimates.

Three linear regression models were used to investigate potential associations with each biomarker and adjusted for potential confounders. Model 1 (‘minimally-adjusted’) adjusted for age, sex, baseline assessment centre, principal components 1-5 for population stratification, and genotyping array. Model 2 (‘partially-adjusted’) also included self-reported diabetes, high blood pressure and coronary heart disease (comprised of angina plus myocardial infarction [22]). Model 3 (‘fully adjusted’) also controlled for self-reported cholesterol, hormone replacement therapy, oral contraceptive, insulin or hypertension medication, Townsend deprivation scores, and ever vs. never smoking. The cross-sectional association between *APOE* vs. dementia outcomes were also analysed using these models.

As additional sensitivity analyses we adjusted for dummy-variable ‘any self-reported chronic inflammatory condition’ (n=64,996; 17%); underweight (BMI<18.5; n=1,962 or 0.5%) or obesity (BMI≥30, n = 297,738 or 75.4%) vs. normal to overweight (18.5 to 30 BMI; n=95,001 or 24.1%), month of assessment, and finally we corrected any significant associations with IGF-1 for self-reported baseline cancer history (n=33,406; 8%) [23]. SNP data was collated and quality controlled with PLINK V1.90, and analysed with Stata V16.

## Results

We excluded participants with non-white British ancestry (N=78,672; 16%), sex mismatch (self-report versus genetic), chromosomal aneuploidy, excessive heterozygosity and genotype missing rate >10%. We excluded the minority of participants with e2/e4 (n=2,556; 0.7%) genotype because this included potentially protective and risk alleles [24]. We removed outliers >5SDs from the mean for each biomarker. This left overall N=395,769 which varied slightly by biomarker: Table 1 shows sample size and key values per biomarker.

**Table 1:**
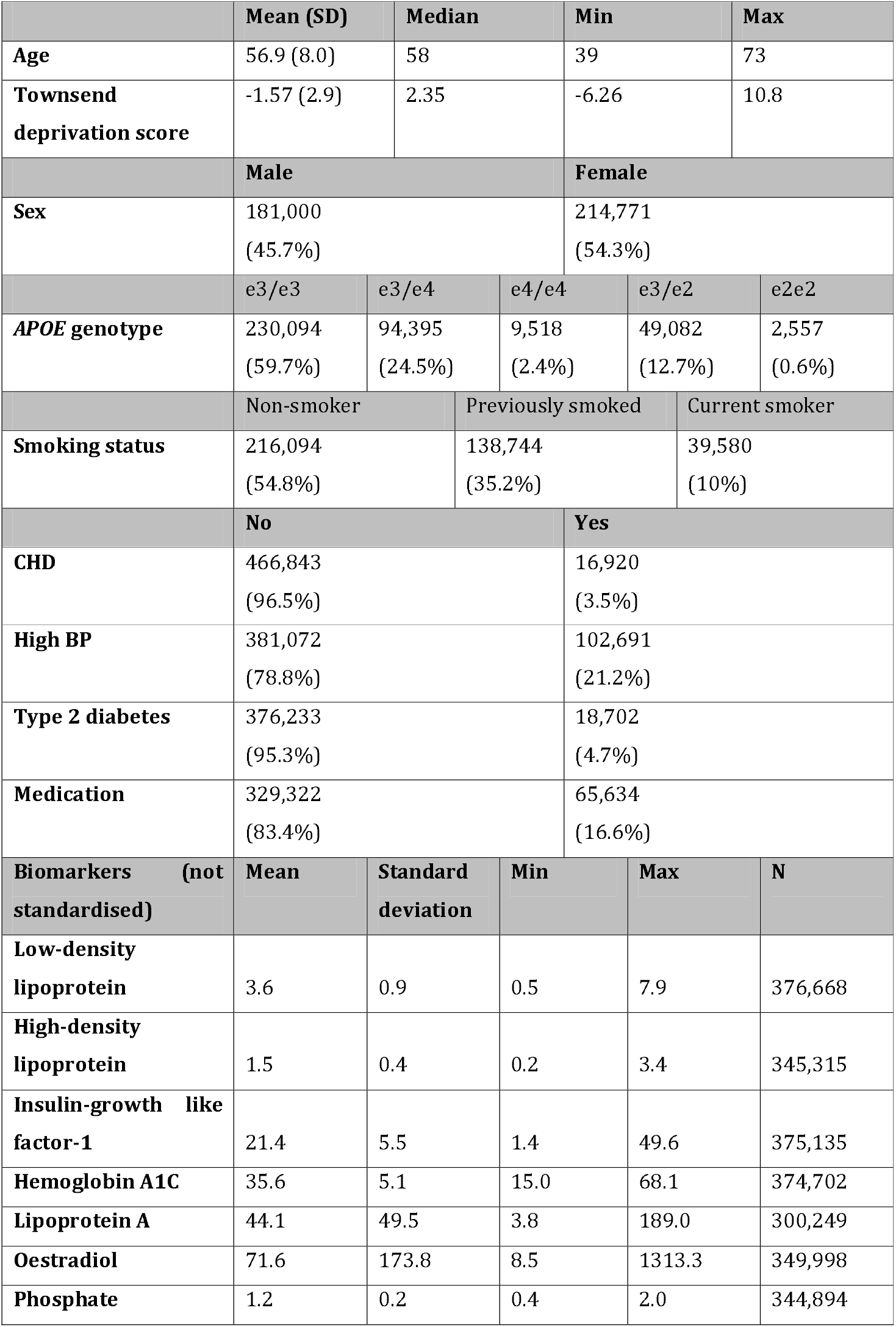
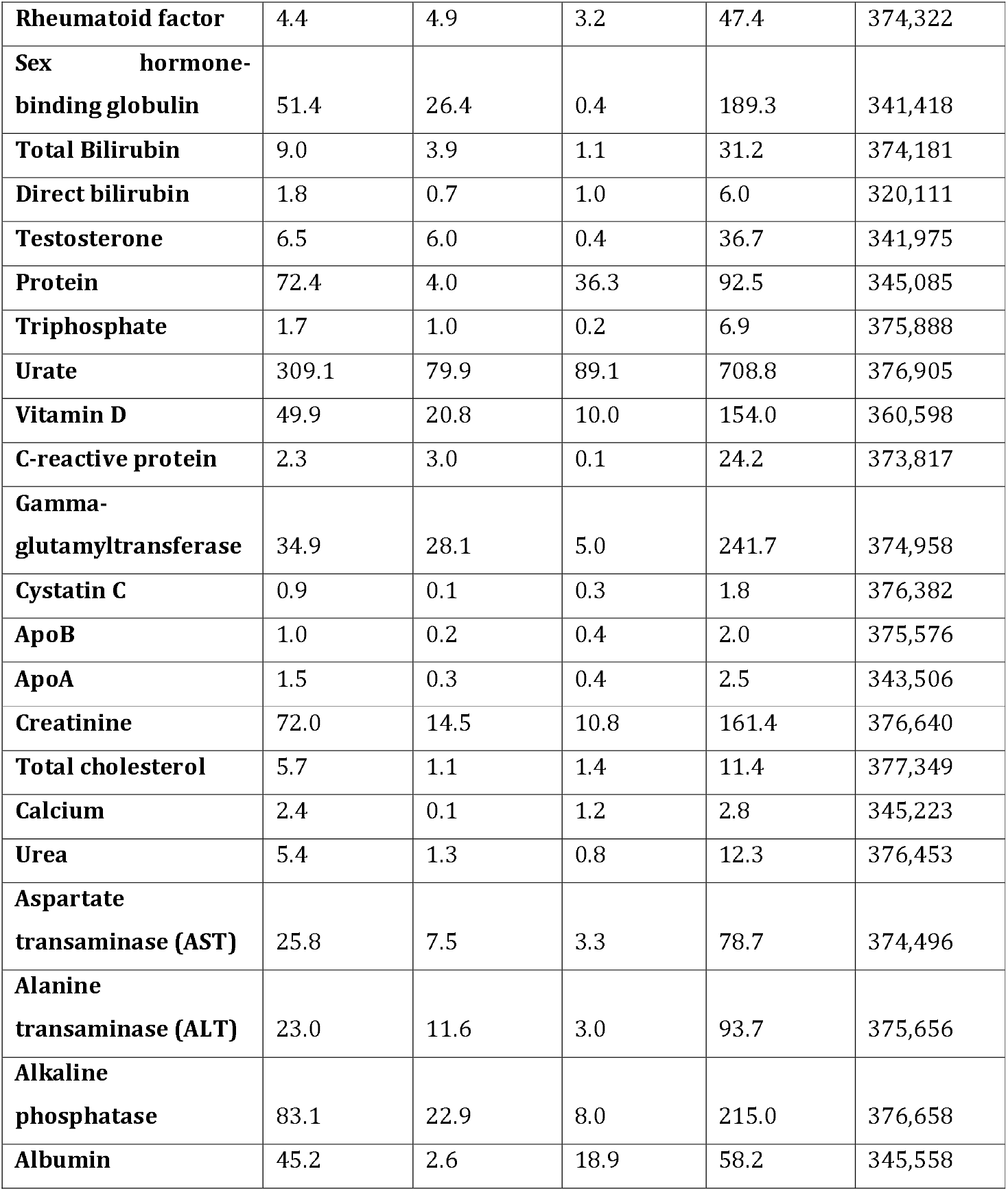
Baseline descriptive values.

|  | Mean (SD) | Median | Min | Max |  |
| --- | --- | --- | --- | --- | --- |
| **Age** | 56.9 (8.0) | 58 | 39 | 73 |  |
| **Townsend deprivation score** | -1.57 (2.9) | 2.35 | -6.26 | 10.8 |  |
|  | Male |  | Female |  |  |
| **Sex** | 181,000<br>(45.7%) |  | 214,771<br>(54.3%) |  |  |
|  | e3/e3 | e3/e4 | e4/e4 | e3/e2 | e2/e2 |
| **APOE genotype** | 230,094<br>(59.7%) | 94,395<br>(24.5%) | 9,518<br>(2.4%) | 49,082<br>(12.7%) | 2,557<br>(0.6%) |
|  | Non-smoker |  | Previously smoked |  | Current smoker |
| **Smoking status** | 216,094<br>(54.8%) |  | 138,744<br>(35.2%) |  | 39,580<br>(10%) |
|  | No |  |  | Yes |  |
| **CHD** | 466,843<br>(96.5%) |  |  | 16,920<br>(3.5%) |  |
| **High BP** | 381,072<br>(78.8%) |  |  | 102,691<br>(21.2%) |  |
| **Type 2 diabetes** | 376,233<br>(95.3%) |  |  | 18,702<br>(4.7%) |  |
| **Medication** | 329,322<br>(83.4%) |  |  | 65,634<br>(16.6%) |  |
| Biomarkers (not standardised) | Mean | Standard deviation | Min | Max | N |
| **Low-density lipoprotein** | 3.6 | 0.9 | 0.5 | 7.9 | 376,668 |
| **High-density lipoprotein** | 1.5 | 0.4 | 0.2 | 3.4 | 345,315 |
| **Insulin-growth like factor-1** | 21.4 | 5.5 | 1.4 | 49.6 | 375,135 |
| **Hemoglobin A1C** | 35.6 | 5.1 | 15.0 | 68.1 | 374,702 |
| **Lipoprotein A** | 44.1 | 49.5 | 3.8 | 189.0 | 300,249 |
| **Oestradiol** | 71.6 | 173.8 | 8.5 | 1313.3 | 349,998 |
| **Phosphate** | 1.2 | 0.2 | 0.4 | 2.0 | 344,894 |
| **Rheumatoid factor** | 4.4 | 4.9 | 3.2 | 47.4 | 374,322 |
| **Sex hormone-binding globulin** | 51.4 | 26.4 | 0.4 | 189.3 | 341,418 |
| **Total Bilirubin** | 9.0 | 3.9 | 1.1 | 31.2 | 374,181 |
| **Direct bilirubin** | 1.8 | 0.7 | 1.0 | 6.0 | 320,111 |
| **Testosterone** | 6.5 | 6.0 | 0.4 | 36.7 | 341,975 |
| **Protein** | 72.4 | 4.0 | 36.3 | 92.5 | 345,085 |
| **Triphosphate** | 1.7 | 1.0 | 0.2 | 6.9 | 375,888 |
| **Urate** | 309.1 | 79.9 | 89.1 | 708.8 | 376,905 |
| **Vitamin D** | 49.9 | 20.8 | 10.0 | 154.0 | 360,598 |
| **C-reactive protein** | 2.3 | 3.0 | 0.1 | 24.2 | 373,817 |
| **Gamma-glutamyltransferase** | 34.9 | 28.1 | 5.0 | 241.7 | 374,958 |
| **Cystatin C** | 0.9 | 0.1 | 0.3 | 1.8 | 376,382 |
| **ApoB** | 1.0 | 0.2 | 0.4 | 2.0 | 375,576 |
| **ApoA** | 1.5 | 0.3 | 0.4 | 2.5 | 343,506 |
| **Creatinine** | 72.0 | 14.5 | 10.8 | 161.4 | 376,640 |
| **Total cholesterol** | 5.7 | 1.1 | 1.4 | 11.4 | 377,349 |
| **Calcium** | 2.4 | 0.1 | 1.2 | 2.8 | 345,223 |
| **Urea** | 5.4 | 1.3 | 0.8 | 12.3 | 376,453 |
| **Aspartate transaminase (AST)** | 25.8 | 7.5 | 3.3 | 78.7 | 374,496 |
| **Alanine transaminase (ALT)** | 23.0 | 11.6 | 3.0 | 93.7 | 375,656 |
| **Alkaline phosphatase** | 83.1 | 22.9 | 8.0 | 215.0 | 376,658 |
| **Albumin** | 45.2 | 2.6 | 18.9 | 58.2 | 345,558 |

### APOE associations with dementia phenotypes

As a form of replication and to support the utility of investigating *APOE* genotype in the UK Biobank cohort, we tested for e4 and e2 allele count (vs. e3/e3) against all-cause dementia (n=1,852; 0.5%), and specific AD diagnosis (n=722; 0.2%). We found that e4 was associated with increased AD (fully-adjusted OR = 3.51 per allele, 95% CI = 3.14 to 3.92, P<0.001) and all-cause dementia (OR = 2.59, 95% CI = 2.40 to 2.79, P<0.001), whereas, e2 was correspondingly associated with decreased AD (OR = 0.59, 95% CI = 0.40 to 0.85, P = 0.005) and all-cause dementia (OR = 0.78, 95% CI = 0.65 to 0.93, P = 0.007). An unadjusted chi-square test showed 64%% of people with AD had at least one e4 allele vs. 36% in the non-AD group.

### APOE associations with biomarker values

Several significant associations (at nominal p<0.05) were identified between *APOE* genotype status and biomarker values, as shown in Figure 1. Increasing e4 allele count associated with significant differences in several biomarker values (Supplementary Table S1). There were associations between e4 allele count vs. higher LDL, IGF-1, sex hormone binding globulin (SHBG), total bilirubin, triphosphate levels, ApoB and total cholesterol. Negative associations were found between e4 genotype and lower high-density lipoprotein (HDL), haemoglobin A1c (HbA1c), lipoprotein A, phosphate, C-reactive protein (CRP), gamma glutamyl transferase (GGT), vitamin D, creatinine, urate, and urea. The largest effect sizes were seen for total cholesterol (0.13 SDs per allele in the fully adjusted model), ApoB (0.20), CRP (−0.12) and LDL (0.15), with the rest <0.1SDs per e4 allele. There was no evidence of a significant fully adjusted association between e4 and oestradiol, aspartate transaminase (AST), albumin, testosterone and rheumatoid factor (RF) levels.

**Figure 1:**
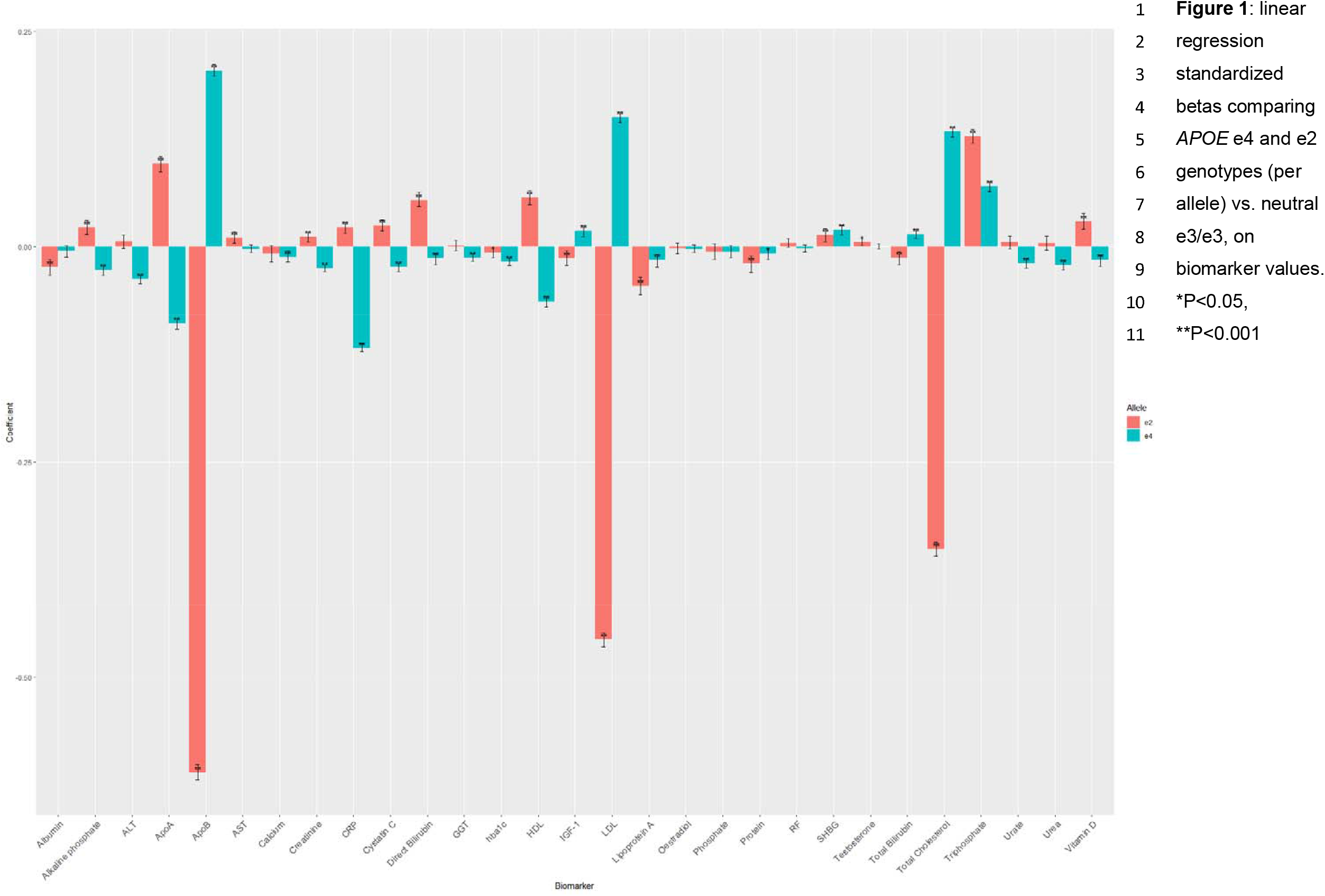
linear regression standardized betas comparing *APOE* e4 and e2 genotypes (per allele) vs. neutral e3/e3, on biomarker values. *P<0.05, **P<0.001.

Significant associations were seen between e2 allele count vs. lower LDL, HDL, IGF-1, total bilirubin, direct bilirubin, vitamin D, CRP, cystatin C (CysC), ApoA, ApoB, creatinine and alkaline phosphatase (Figure 1). These are in the opposite directions of effect reported for the associations to the e4 allele (vs. e3/e3) as expected. Associations between *APOE* e2 and HbA1c, lipoprotein A, SHBG and triphosphate levels were also identified, however, these effects were in the same direction as e4. The largest effect sizes were for LDL (−0.46 SDs per allele in the fully-adjusted model), triphosphate (0.13), ApoB (−0.61) and total cholesterol (−0.35), with the rest <0.1SDs per e2 allele.

There were instances of significant association for e2 but not e4 vs. e3/e3. These were: negative associations between e2 vs. protein and aspartate aminotransferase levels, and positive association between e2 and testosterone. There was no statistically significant association between e2 count and oestradiol, phosphate, GGT, urate, urea and alanine aminotransferase levels (Supplementary Table S1). Both e4 and e2 were also found to associate with increased SHBG, decreased protein and increased triphosphate (vs. e3/e3).

### Sex-specific analyses

There were instances of male/female sex vs. genotype interactions for several biomarkers (Supplementary Table S2). Out of 33 biomarkers, 16 showed some interaction (P<0.05): LDL, HDL, HbA1c, Oestradiol, RF, SHBG, Testosterone, protein, triphosphate, urate, ApoB, ApoA, total cholesterol, AST and alanine transaminase (ALT). We provide complete sex-specific Z-score associations in Supplementary Table S3. Most of the significant associations in the collated analyses remained so in individual sexes. Certain associations were only significant in males: e4 vs. oestradiol and calcium, and e2 vs. IGF-1, SHBG, testosterone and cystatin c.

### Sensitivity analyses

All nominally significant associations survived correction for FDR. Results were unchanged in terms of effect size and P-value when individuals with incident dementia/AD were removed from analyses. When we added presence of any self-reported inflammatory condition, month of assessment, underweight or obesity (vs. normal to overweight) to the final model, no results were meaningfully changed. IGF-1 results were unchanged when we additionally controlled for all-cause cancer. As a check, we re-ran all final-model tests as 0 vs. 1 allele, and 0 vs. 2 allele contrasts (rather than a 0/1/2 dose effect); results were consistently indicative of dose effects in the same direction.

## Discussion

The *APOE* e4 allele is known to associate in UK Biobank with worse non-demented cognitive abilities [17], cerebrovascular health [3] and here clinically-ascertained AD/dementia risk. In this study we found several significant associations per e4 allele on circulating biomarker levels (vs. neutral e3/e3). In many instances these were supported by corresponding associations between the putatively protective e2 genotype in the opposite direction as would be expected. Gaining a greater understanding of the biomarker profiles of individuals at genetic risk of AD may be useful in the future for earlier detection of AD and could potentially highlight pathways as therapeutic targets [11]. In some instances, the directions of effect for e4 (‘risk’) and e2 (‘protective’) alleles conflicted with what would be expected based on levels reported elsewhere in people with prevalent AD. For example: lower levels of IGF-1 have previously associated with increased risk of AD and cognitive decline [6,25]. By contrast here we showed association between e4 and increased IGF-1, and between e2 genotype and lower IGF-1. This is surprising as IGF-1 stimulates neurogenesis and promotes cell survival [26]. Previous reports could reflect some degree of reverse causality or bias in cross-sectional AD patient sample studies showing lower IGF-1 levels i.e. where disease onset affects biomarker health.

### Interpretation

Previous studies have reported *APOE* e4 association with higher vitamin D serum concentration [27] however here e4 associated with decreased, and e2 with increased vitamin D. In e4 homozygotes, a higher vitamin D concentration has been associated with higher memory function, suggesting higher vitamin D levels could be protective for people at risk for AD [28].

Investigations into the effect of differing cholesterol levels on cognitive decline and AD have produced conflicting results with both low and high cholesterol being associated [8,29– 31]. The lack of consensus could be partially due to the smaller sample sizes previously used. The three *APOE* alleles encode for different ApoE protein isoforms with altered lipid interactions in serum; the e3 encoded protein isoform is associated with “normal” plasma lipid levels [32]. *APOE* e4 and its associated higher LDL have been previously associated with early onset of AD [33] while e2 has protective effects on the concentrations of cholesterol, lipids and phospholipids [34]. Here we reinforce those observations [5,8,34,35].

It has been hypothesised that lipid metabolism is important in the pathophysiology of AD [11]. The significant associations between *APOE* genotype status and ApoA/ApoB support this. ApoA and ApoB proteins are major surface proteins of HDL and LDL, respectively [36]; previously ApoA has been associated with lower risk of cardiovascular disease [37], whereas, ApoB is reported to be proatherogenic [36]. We identified lower ApoA levels in e4 carriers and higher levels in e2 carriers; ApoA has reportedly neuroprotective effects by inhibiting β-amyloid plaque aggregation [36,38]. Our findings are supported by previous associations reported between decreased serum ApoA and increased AD risk [38]. With ApoB, we found e4 was associated with elevated levels of ApoB and e2 with lower levels of ApoB compared to e3. This is consistent with the direction of effect reported in most [10,33] but not all studies [36].

It is hypothesised *APOE* allele-encoded protein isoforms have different affinities for lipoproteins [39]. It has been difficult to define the exact effects of *APOE* genotype on lipoprotein A from previous studies; many previous studies were limited by small sample sizes. Our data showed that both *APOE* e2 and e4 were significantly associated with decreased levels of lipoprotein A compared to e3. Another relatively large study (N=46,615) has also reported evidence of *APOE* e2 and e4 carriers displaying lower lipoprotein A levels [39]. Elevated levels of lipoprotein A is are known to have a causal relationship with myocardial infarction, which has previously been associated with increased dementia risk [39]. The influence of lower lipoprotein A on the pathophysiology of AD remains unclear.

It has been suggested that *APOE* genotype status modifies the effect of sex hormones on AD and dementia symptoms [40]. We did not find any evidence for a significant association between *APOE* genotype and oestradiol in the whole sample, although in males e4 associated with lower levels. Oestradiol has suggested neuroprotective effects and sex-specific effects on AD patients; however, evidence is conflicting, and this relationship is not fully understood [40]. There was an association found between *APOE* e2 and higher testosterone levels, specific to males. There is conflicting evidence regarding the effect of testosterone on AD risk [41,42]; some cross-sectional studies report lower levels of testosterone are more often observed in AD patients compared to healthy controls [42] which would be consistent with our findings as e2 is associated with decreased AD risk [1].

The underlying pathophysiology of AD has been suggested to be at least partially influenced by neuroinflammation [43]. Serum CRP levels are a marker for inflammation but the evidence for association with AD risk is conflicting. Interactions between *APOE* e4 and elevated CRP have been reported to associate with early onset of AD [44]. However, consistent with our findings e4 has been associated with lower CRP levels, and e2 with higher CRP levels [10,45]. Further research is required to elucidate the effects of CRP levels on the pathophysiology of AD, particularly given it is a marker of acute rather than chronic inflammation [43]. Another inflammatory marker associated with increased risk of AD is raised GGT [46]. We identified lower levels of GGT in e4 carriers: this may reflect bias in cross-sectional studies of GGT and AD.

Alkaline phosphatase may have some involvement in the inflammatory/AD process [47].We identified lower levels of alkaline phosphatase in e4 carriers and elevated levels in e2 carriers. It has been suggested that alkaline phosphatase could potentially be used as a therapy to reduce neuroinflammation in AD [47]; our findings may support this but more in depth investigations are required.

CysC, typically a marker of kidney dysfunction, is also involved in modulation of inflammatory responses and reported to have neuroprotective effects in AD as it co-localizes with β-amyloid and inhibits oligomerization [48]. We found that e4 associated with lower CysC and e2 with higher CysC levels compared to e3. Lower baseline CysC has been reported to precede AD onset in an 11-year longitudinal study of non-demented elderly men at baseline; in one small study (N=82) [49] which suggested the finding may be due to attrition bias because higher CysC is a risk factor for cardiovascular disease and earlier mortality. Our findings potentially lend support to low CysC serum levels as an AD risk factor.

We report suggestive association between e4 and lower phosphate levels, potentially contradicting prior research showing association between (age-dependent) higher serum phosphorous and incident dementia [50]. We found significant associations between e4 and higher total bilirubin and urea, and lower direct bilirubin, urate, creatinine, calcium, and alanine aminotransferase. In the whole sample analysis, we found evidence of associations between e2 and higher direct bilirubin, creatinine and aspartate aminotransferase, and lower total bilirubin and albumin. Both e4 and e2 associated in same direction with HbA1c, SHBG, protein and triphosphate. This is unexpected: e4 and e2 tend to show opposing effects regarding AD. We found proportionally moderate evidence of interaction between sex, genotype and biomarker; this is a significant area of research and warrants further study [12].

### Limitations and future research

A limitation of UK Biobank is that participants are overall likely to have fewer health conditions, be better educated, of older age, female, and living in less socio-economically deprived areas than the general population [13]. This study was conducted in individuals of White European ancestry only and so these results may not be generalizable to a more mixed population. Findings may not be truly representative of the effects of *APOE* genotype in the wider UK population [13]. There may be conflicting biases at play in at least some of the results. For example, confounding effects may exist where e4 carriers are of poorer health, affecting their lifestyle and in turn influencing biomarker values, or by contrast selection bias, where the e4 carriers here are of particularly good health relative to the general population.

Although this study reports significant associations between serum biomarkers and *APOE* genotype, which could influence the risk of AD, these alleles are unlikely to be entirely responsible [1]. Some biomarker levels may only be pathogenic in combination with other biomarkers [8]. Some of the biomarkers may be influenced by environmental factors such as seasonality, although this should to some extent average out; sensitivity analyses showed no evidence of confounding. The effect sizes reported here are in many cases small and the results require replication; the clinical applicability of these findings remains unclear. It is not necessarily possible to identify the exact biological pathways involved in the pathophysiology of AD from these analyses. Further work is required to investigate the underlying pathways to identify processes which could be modified or targeted to decrease the risk of AD, including longitudinal biomarker data [12,35]. Future research e.g. using Mendelian randomization may investigate whether pharmaceutically altering serum biomarker levels or implementing lifestyle changes to manage these biomarkers may be beneficial to individuals at greater risk of developing AD.

## Conclusion

The exact influence of *APOE* on AD and dementia pathophysiology is unclear. Through this study we have identified associations between the high-risk AD gene locus *APOE* and a range of serum blood biomarkers in UK Biobank. These associations highlight potential pathways involved in the development of cognitive impairment and have potential to lead to earlier detection of AD risk through the analysis of biomarkers and *APOE* genotype.

## Data Availability

There are restrictions prohibiting the provision of data in this manuscript. The data were obtained from a third party, UK Biobank, upon application. Interested parties can apply for data from UK Biobank directly, at http://www.ukbiobank.ac.uk. UK Biobank will consider data applications from bona fide researchers for health-related research that is in the public interest. By accessing data from UK Biobank, readers will be obtaining it in the same manner as we did.

## Acknowledgements

This research has been conducted using the UK Biobank resource; we are grateful to UK Biobank participants. This project was completed using UK Biobank application 17689 (PI: DML).

## Author contributions

Study concept and design: DML.

Statistical analysis: ACF; DML.

Drafted the manuscript: ACF, DML.

Critically revised content: all authors.

Obtained principal study funding: DML.

